# Comparable in vivo joint kinematics between self-reported stable and unstable knees after TKA can be explained by muscular adaptation strategies: a retrospective observational study

**DOI:** 10.1101/2022.12.12.22283339

**Authors:** Longfeng Rao, Nils Horn, Nadja Meister, Stefan Preiss, William R. Taylor, Alessandro Santuz, Pascal Schütz

## Abstract

**Background:** Postoperative knee instability is one of the major reasons accounting for unsatisfactory outcomes, as well as a major failure mechanism leading to total knee arthroplasty (TKA) revision. Nevertheless, subjective knee instability is not well defined clinically, plausibly because the relationships between instability and implant kinematics during functional activities of daily living remain unclear. Although muscles play a critical role in supporting the dynamic stability of the knee joint, the influence of joint instability on muscle synergy patterns is poorly understood. Therefore, this study aimed to understand the impact of self-reported joint instability on tibiofemoral kinematics and muscle synergy patterns after TKA during functional gait activities of daily living.

**Methods:** Tibiofemoral kinematics and muscle synergy patterns were examined during level walking, downhill walking, and stair descent in eight self-reported unstable knees after TKA (3M:5F, 68.9±8.3 years, BMI 26.1±3.2 kg/m^2^, 31.9±20.4 months postoperatively), and compared against ten stable TKA knees (7M:3F, 62.6±6.8 years, 33.9±8.5 months postoperatively, BMI 29.4±4.8 kg/m^2^). For each knee joint, clinical assessments of postoperative outcome were performed, while joint kinematics were evaluated using moving video-fluoroscopy, and muscle synergy patterns were recorded using electromyography.

**Results:** Our results reveal that average condylar A-P translations, rotations, as well as their ranges of motion were comparable between stable and unstable groups. However, the unstable group exhibited more heterogeneous muscle synergy patterns and prolonged activation of knee flexors compared to the stable group. In addition, subjects who reported instability events during measurement showed distinct, subject-specific tibiofemoral kinematic patterns in the early/mid-swing phase of gait.

**Conclusions:** Our findings suggest that accurate movement analysis is sensitive for detecting acute instability events, but might be less robust in identifying general joint instability. Conversely, muscle synergy patterns seem to be able to identify muscular adaptation associated with underlying chronic knee instability.

**Funding:** This research received no specific grant from any funding agency in the public, commercial, or not-for-profit sectors.

## Introduction

Postoperative knee instability is one of the major reasons accounting for unsatisfactory outcomes, as well as a major failure mechanism leading to revision surgery after primary total knee arthroplasty (TKA). However, postoperative knee instability is not well defined clinically, and the boundaries between stable and unstable TKA knees is still poorly understood. This lack of understanding is likely due to the multitude of factors that are associated knee instability, including inadequate soft tissue balancing (1, 2), loss of ligamentous integrity (3), and improper component sizing (3, 4).

Clinical assessment of joint stability relies on passive laxity examinations and patient reported outcome measures (PROMs). Manual stress tests including the anterior/posterior drawer, Lachman, and varus/valgus tests, as well as questionnaires such as the Knee injury and Osteoarthritis Outcome Score (KOOS) (5), Oxford Knee Score (OKS) (6), University of California and Los Angeles (UCLA) Activity Scale (7), and the Western Ontario and McMaster Universities Osteoarthritis Index (WOMAC) (8) are widely used in clinical practice to assess knee instability after TKA. However, such clinical assessments are highly subjective with large inter-rater variability (9). With the goal to objectively investigate knee instability, methods for quantifying passive knee laxity such as the KT-1000/2000 (10, 11), Telos (12-14), Rotometer (15, 16), and Rolimeter (17) devices, all generally combined with stress radiography, have been presented. Studies using these techniques have demonstrated that passive knee laxity plays a key role for functional outcomes and patient satisfaction after TKA (18, 19). Nevertheless, it remains unclear whether self-reported instability after TKA is reflected in implant kinematics *in vivo* during functional activities of daily living. Specifically, downhill walking and stair descent are considered as challenging tasks for subjects with knee pathologies (20, 21), and could therefore present relevant activities for provoking feelings of instability.

As the primary actuators of the locomotion system, muscles play a critical role in guiding motor functionality as well as supporting stability of the knee joint. Muscular deficits are often observed after TKA and closely correlate with physical impairment and limitations in daily activities (22, 23). Well-coordinated muscular activity is thought to be necessary for normal gait patterns (22). Here, adaptation of muscle recruitment patterns in the early postoperative phase, where increased knee extensor (quadriceps) and flexor (hamstrings) co-activation has generally been observed, and is thought to enhance knee stability (24, 25). However, whether such muscular adaptation is temporary and can be reversed in the long-term after TKA remains controversially discussed (25, 26). Moreover, it remains unknown whether such adaptive strategies persist in unstable knees, or whether different lower limb muscle synergy patterns develop to compensate for deficits in knee stability.

While the accurate assessment of tibiofemoral kinematics during functional activities is challenging, the recent development of dynamic video-fluoroscopy systems now provides access to implant kinematics with a high level of accuracy and without soft tissue artefact throughout consecutive cycles of gait activities (27-30). Such systems have recently allowed investigations into the impact of activity (29) and implant geometry (28, 30) on tibiofemoral kinematics during functional activities such as level walking, downhill walking, and stair descent, and can be combined with electromyography (EMG) to assess muscle activation synergies and its role on modulating joint kinematics (25, 27, 31). As such, the application of these approaches to subjects with stable and unstable knees after TKA could establish whether compensation mechanisms continue to occur, but also elucidate the role of neuromuscular activation and coordination on kinematic adaptations in the joint.

To understand tibiofemoral kinematics in unstable TKA knees and its interaction with muscle activation strategies, the objective of this study was to investigate *in vivo* tibiofemoral implant kinematics and muscle synergy patterns in subjects with stable and self-reported unstable knees after TKA during functional activities of daily living using dynamic video-fluoroscopy and EMG. Specifically, we aimed to establish whether unstable knees exhibit higher levels of relative tibiofemoral motion and distinct muscle synergy patterns/strategies than their stable counterparts.

## Methods

### Study cohorts

In total, 17 subjects (age ≥45 years, pain VAS≤3) with 18 replaced knees implanted with Persona cruciate retaining (CR) TKA components and an ultra-congruent (UC) inlay (Zimmer Biomet, Warsaw, IN, USA) were recruited at least one year postoperatively. Both cruciate ligaments were sacrificed in all TKA knees. Of the 18, 8 TKA knees (3M:5F, 68.9±8.3 years, 31.9±20.4 months postoperatively, BMI 26.1±3.2 kg/m^2^) were recruited from subjects reporting episodes of buckling, shifting, or giving away of the operated knee during daily activities in the three months prior to recruitment, with or without clinical signs of instability (unstable cohort). Ten TKA knees were recruited into the stable group (7M:3F, 62.6±6.8 years, 33.9±8.5 months postoperatively, BMI 29.4±4.8 kg/m^2^) from subjects with no sensation of instability in the operated knee in the same three-month period prior to recruitment. Subjects with significant problems of the lower extremities other than knee instability were excluded from participation in this study, as well as those presenting low back pain, neurological problems, patellofemoral symptoms, aseptic loosening, collateral ligament reconstruction, or an inability to perform the motion tasks, understand and/or sign the informed consent form.

The project was approved by the Zürich cantonal ethics committee (BASEC no. 2019-01242) and all subjects provided their written informed consent prior to participation.

### Clinical assessment

For each subject, clinical assessment of postoperative outcome was performed at the Schulthess Clinic Zürich, including manual passive laxity tests (anterior/posterior drawer and Lachmann, varus/valgus stress, and sagittal passive range of motion), as well as PROMs (OKS, COMI-Knee, EQ-5D-5L and UCLA activity score).

### Experimental procedure

Level gait (straight ahead on a level floor), downhill walking (10° inclined slope), and stair descent (three 18cm steps) were radiographically imaged at 30Hz using the single plane ETH Moving Fluoroscope (32), which allowed *in vivo* tibiofemoral kinematics to be captured throughout complete cycles of each activity. Measurement protocols for the three activities have been described previously (29, 30), but are briefly described here: Prior to each motion task, trials without fluoroscopic imaging were performed until the subject felt comfortable walking with the Moving Fluoroscope. For all activities, 3 to 5 valid cycles (heel-strike to heel-strike for gait activities) were measured. Heel-strike detection was performed using eight force plates (Kistler AG, Winterthur, Switzerland, 2000Hz, force threshold: 25N) or heel marker trajectories (Vicon MX system; Oxford Metrics Group, Oxford, UK; 200Hz) where no force plates were available. Muscular activations were recorded using a wireless 16-channel surface EMG system (Trigno, Delsys, USA) throughout all activities. EMG electrodes were placed on 8 muscles of the TKA limb: rectus femoris, vastus medialis, vastus lateralis, semitendinosus, biceps femoris, tibialis anterior, gastrocnemius medialis, and gastrocnemius lateralis. All measurement systems were temporally synchronized. For confirmation of previous clinical assessments, subjects were asked to report any feelings of instability on a four-point scale after each activity.

### Data processing

Fluoroscopic images were distortion corrected (33) before 2D→3D registration was performed to determine the 3D poses of the implant components for each frame of the activity cycles using an in-house registration software (34) (registration errors: <1° for all rotations, <1mm for in-plane, and <3mm for out of plane translations (32, 33)). To describe relative tibiofemoral rotations, the joint coordinate system approach reported by Grood and Suntay (35) was used based on the local coordinate systems of the respective implant components. Tibiofemoral condylar A-P translations were defined based on the movement of the weighted mean location of the ten nearest points on each condyle relative to a plane on the tibial plateau (29, 30).

Kinematic parameters including tibiofemoral A-P translations, flexion/extension, ab/adduction, and internal/external rotation angles during all gait activities were interpolated to 101 datapoints per cycle. Condylar A-P translations were presented relative to the corresponding medial condyle position at initial heel strike of each trial. For each task, the range of motion (RoM) of all kinematic parameters was determined and separated into stance and swing phases. The intercondylar A-P RoM (lateral RoM – medial RoM) were then used to evaluate the transverse plane pivot location.

EMG data was processed in R (version 4.2.0, R Foundation for Statistical Computing, Vienna, Austria) using package “musclesyneRgies v1.2.5” (36). All EMG signals were filtered (high-pass, cut-off frequency 50Hz, 4^th^ order; full wave rectified; low-pass, cut-off frequency 20Hz, 4^th^ order), amplitude normalized to the maximum value in each subject activity (37), and time-normalized to 200 datapoints, assigning 100 points to the stance and 100 to the swing phases (38). This time-normalisation approach was chosen to ensure that the results could be interpreted independently of the absolute duration of gait events. Muscle synergy weights (time-independent coefficients) as well as the corresponding activation patterns (time-dependent coefficients) were extracted using a non-negative matrix factorization (NMF) algorithm and subsequently functionally classified using k-mean clustering to evaluate consistency of muscle synergies across each group (stable vs. unstable) and activity (total of 6 classifications). Here, the number of classified muscle synergies in each group was imposed based on the average number of muscle synergies extracted per activity (4 for level gait and downhill walking, 3 for stair descent). The full width at half maximum (FWHM) and centre of activity (CoA) of each classified synergy activation pattern were further evaluated and compared between the two groups. The centre of activity, employed to estimate the timing of main activation, was calculated as the angle of the vector in polar coordinates that points to the centre of mass of the circular distribution defined between 0 and 360° or, in other words, between one touchdown and the next (39).

### Statistics

Statistical analysis was performed using MATLAB (R2022a, MathWorks, Natick, MA, USA) and R (version 4.2.0). Independent sample t-tests were conducted to compare differences in tibiofemoral A-P translation RoMs, rotation RoMs, as well as FWHM and CoA of corresponding classified activation patterns between groups. One-dimensional statistical parametric mapping (SPM) (40) was used to test the effects of knee instability on all kinematic parameters over the time series of a gait cycle. One-way ANOVAs were used to examine the effects of knee instability on the muscle synergy weights that demonstrate significant differences in FWHM/CoA of corresponding activation patterns between the groups. For any parameters that exhibit statistical significance between the two groups, corresponding Cohen ’s d effect size (ES) was determined. Statistical significance was set to p<0.05.

## Results

### Clinical assessment

Seven out of the eight unstable knees presented mild hyperextension, while all unstable knees exhibited signs of increased passive laxity for all manual examinations. Only one stable knee, but all unstable knees showed hyperextension. One stable knee and 5 out of 8 unstable knees exhibited increased passive sagittal and coronal laxity. No other differences in clinical examination were observed between the two groups. Inferior OKS and COMI-Knee scores were observed in the unstable knees, even though comparable UCLA activity and EQ-VAS scores were observed between the two groups (Table 1).

**Table 1:** Clinical assessment data of the stable and unstable groups shown as mean ± standard deviation of each parameter. BMI: Body-Mass Index; PTS: Posterior Tibial Slope; RoM: Range of Motion; OKS: Oxford Knee Score; COMI-Knee: Core Outcome Measures Index-Knee; EQ-VAS: EQ-Visual Analogue Scales. Bold values indicate a significant difference.

| Baseline data | Stable (N=10) | Unstable (N=8) | P (2-tailed) |
| --- | --- | --- | --- |
| Age [years] | 62.6 ± 6.8 | 68.9 ± 8.3 | 0.096 |
| BMI [kg/m<sup>2</sup>] | 29.4 ± 4.8 | 26.1 ± 3.2 | 0.113 |
| Time post-op [months] | 33.9 ± 8.5 | 31.9 ± 20.4 | 0.779 |
| PTS [°] | 82.0 ± 2.4 | 82.3 ± 3.0 | 0.863 |
| Inlay thickness [mm] | 11.3 ± 1.0 | 11.8 ± 1.7 | 0.511 |
| Knee flexion RoM [°] | 125.0 ± 7.8 | 126.3 ± 6.4 | 0.720 |
| Hypertension | 1/10 | 8/8 | \ |
| Drawer tests | 1/10 | 5/8 | \ |
| Varus/valgus tress tests | 1/10 | 5/8 | \ |
| UCLA activity score | 8.3 ± 1.3 | 7.9 ± 1.1 | 0.465 |
| OKS | 46.0 ± 2.0 | 42.9 ± 3.8 | **0.040** |
| COMI-Knee | 0.2 ± 0.6 | 0.9 ± 0.8 | **0.044** |
| EQ-VAS | 84.5 ± 13.4 | 81.3 ± 17.1 | 0.660 |

### A-P translations

Video-fluoroscopic analysis of the functional tibiofemoral kinematics revealed comparable mean A-P translation patterns for both the medial and lateral condyles between the stable and unstable groups throughout the stance and swing phases of all activities (Figure 1). For level and downhill walking, variability was generally higher in unstable TKA knees than their stable counterparts. Interestingly, however, unstable knees exhibited less variability in medial condylar A-P translation during late-stance and early-swing phase (≈ 60-75% gait cycle) compared to stable knees.

**Figure 1:**
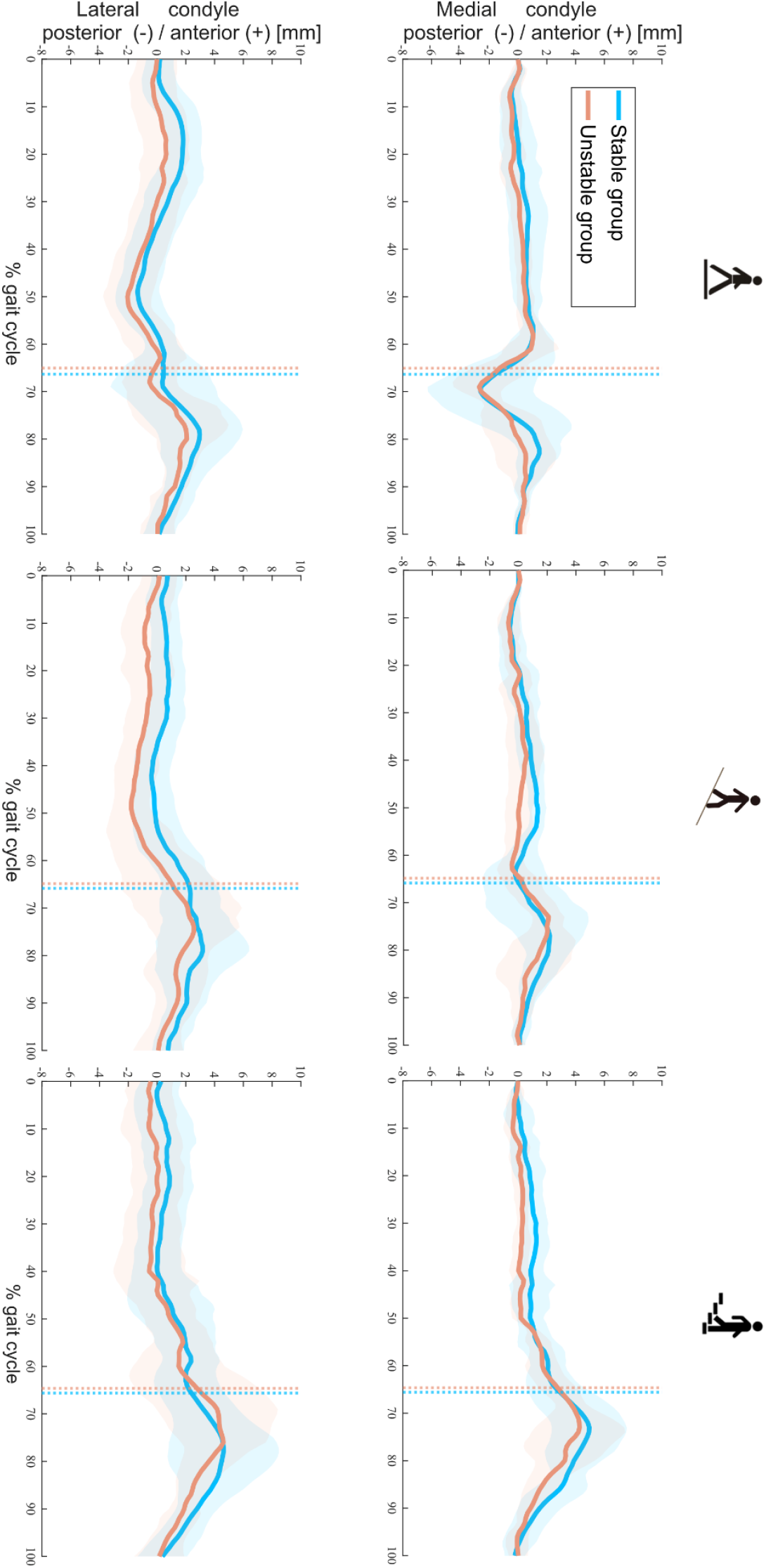
Tibiofemoral A-P translations in stable and unstable TKA knees during level walking (left), downhill walking (middle), and stair descent (right). Means (solid lines) and standard deviation (shaded areas) of A-P translations in both groups are presented. Dotted colour lines indicate the mean toe-offs for the stable and unstable groups.

The ranges of medial and lateral condylar A-P translations showed comparable results between the stable and unstable groups throughout all activities (Table 2). However, the inter-condylar differences in A-P translations demonstrated significant differences between groups during the stance phase of downhill walking (unstable 1.1±1.3mm, stable -0.2±1mm, p = 0.04) and the swing phase of stair descent (unstable 0.5±2.4mm, stable -1.7±2.0mm, p = 0.02).

**Table 2:** Mean ± standard deviation of the A-P range of motion (RoM) for the medial and lateral condyles as well as differences between condylar ranges of motion (Diff), calculated as lateral minus medial A-P translation, are reported for stance and swing phases during level walking, downhill walking, and stair descent. Significant differences between stable and unstable groups are indicated using ^a^ and ^b^.

| A-P translation RoM [mm] |  | Stance phase |  |  | Swing phase |  |  |
| --- | --- | --- | --- | --- | --- | --- | --- |
|  |  | Medial | Lateral | Diff | Medial | Lateral | Diff |
| Level walking | stable | 5.4±1.4 | 5.1±1.0 | -0.3±2.2 | 7.0±2.5 | 5.7±1.9 | -1.1±1.5 |
|  | unstable | 4.5±0.9 | 4.5±0.8 | -0.3±1.8 | 6.2±1.6 | 5.5±1.5 | -0.6±2.1 |
| Downhill walking | stable | 4.2±0.9 | 4.0±0.7 | -0.2±1.0<sup>a</sup> | 5.7±1.5 | 4.7±1.2 | -1.0±1.6 |
|  | unstable | 3.4±1.0 | 4.5±0.7 | 1.1±1.3<sup>a</sup> | 5.5±1.5 | 5.4±2.0 | -0.1±1.6 |
| Stair descent | stable | 5.1±1.6 | 5.5±1.9 | 0.4±0.8 | 7.7±1.7 | 6.0±2.1 | -1.7±2.0<sup>b</sup> |
|  | unstable | 4.6±1.2 | 5.5±1.7 | 0.9±0.8 | 6.9±1.7 | 7.4±2.9 | 0.5±2.4<sup>b</sup> |

### Rotations

No significant differences in knee rotations were observed between the two groups during either the stance or swing phase for all tasks (Figure 2). Only the range of ab/adduction during the stance phase of downhill walking exhibited a significant difference between the cohorts (unstable 2.8±0.3°, stable 2.2±0.3°, p<0.001, Table 3).

**Table 3:** Mean ± Standard deviation of knee range of flexion/extension (flex/ex), abduction/adduction (ab/ad), and internal/external (int/ext) rotations for the stance and swing phases of level walking, downhill walking, and stair descent. A significant difference was observed only between stable and unstable groups in abduction/adduction during downhill walking (^a^).

| Rotation RoM [°] |  | Stance phase |  |  | Swing phase |  |  |
| --- | --- | --- | --- | --- | --- | --- | --- |
|  |  | flex/ex | int/ext | ab/ad | flex/ex | int/ext | ab/ad |
| Level walking | stable | 47.2±4.9 | 7.7±1.1 | 2.1±0.2 | 61.8±4.4 | 6.9±1.8 | 2.8±0.5 |
|  | unstable | 43.5±6.8 | 7.0±1.8 | 2.1±0.3 | 59.6±8.0 | 7.8±2.5 | 2.7±0.7 |
| Downhill walking | stable | 57.9±3.8 | 5.5±0.9 | 2.2±0.3<sup>a</sup> | 68.6±4.6 | 7.0±1.3 | 2.5±0.4 |
|  | unstable | 54.9±4.3 | 6.1±1.2 | 2.8±0.3<sup>a</sup> | 67.4±6.7 | 8.2±1.8 | 2.8±0.7 |
| Stair descent | stable | 84.4±3.4 | 8.7±1.2 | 3.5±0.5 | 91.2±4.5 | 9.4±1.9 | 2.8±0.6 |
|  | unstable | 84.5±11.7 | 8.4±1.0 | 3.3±0.4 | 92.8±5.8 | 10.3±2.6 | 3.1±0.9 |

**Figure 2:**
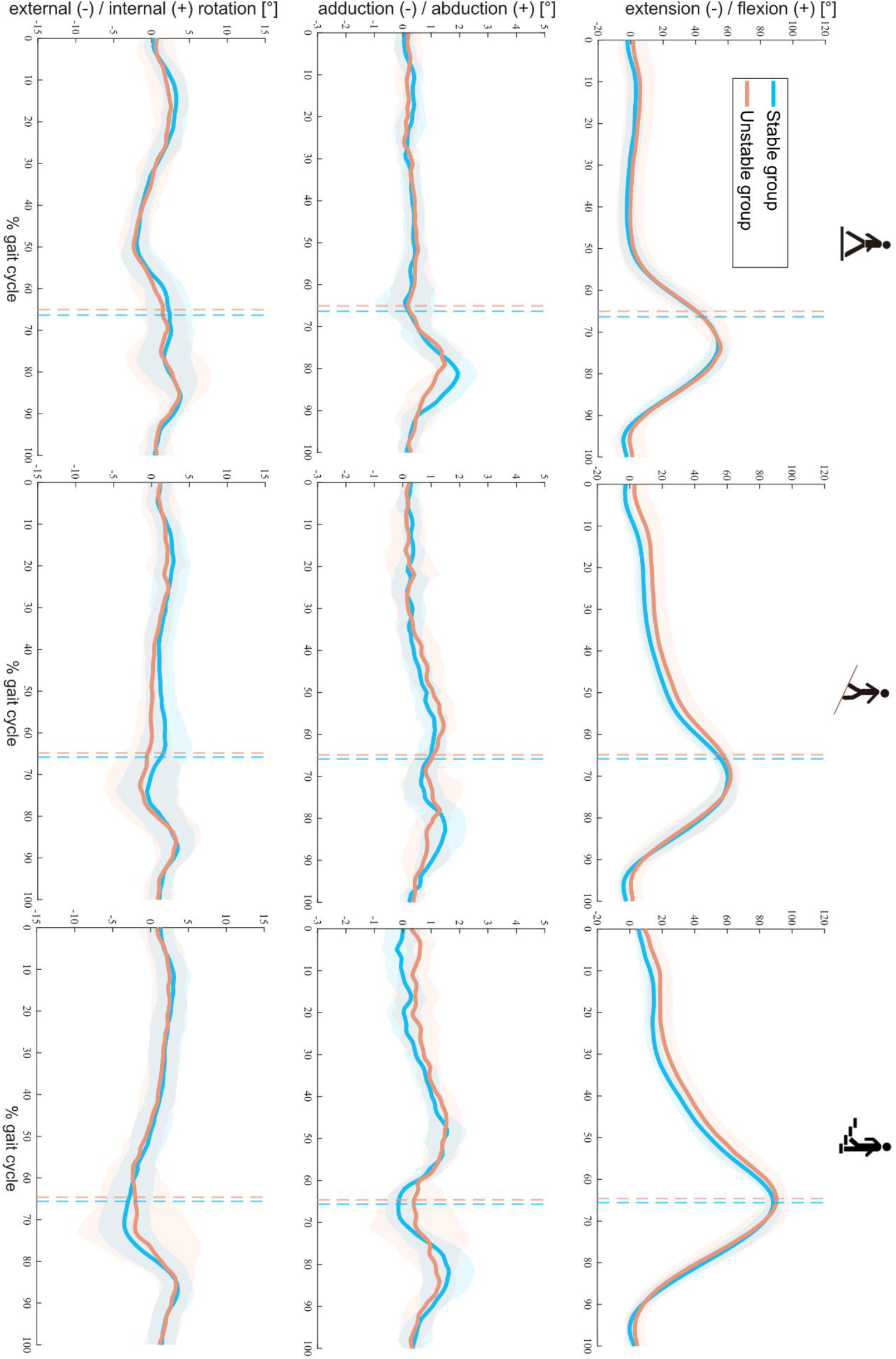
Tibiofemoral rotations throughout a gait cycle in stable and unstable TKA knees during level walking (left), downhill walking (middle), and stair descent (right). Means (solid lines) and standard deviations (shaded areas) of both groups are presented. Dotted lines indicate the mean toe-offs for each group.

### Instability during activity

Within the unstable group, 3 subjects specifically reported the feeling of joint instability while undertaking the measured activities. Interestingly, all 3 subjects reported instability during the more challenging activities of downhill walking and stair descent, where individual kinematic outlying characteristics were observable (Figure 3).

**Figure 3:**
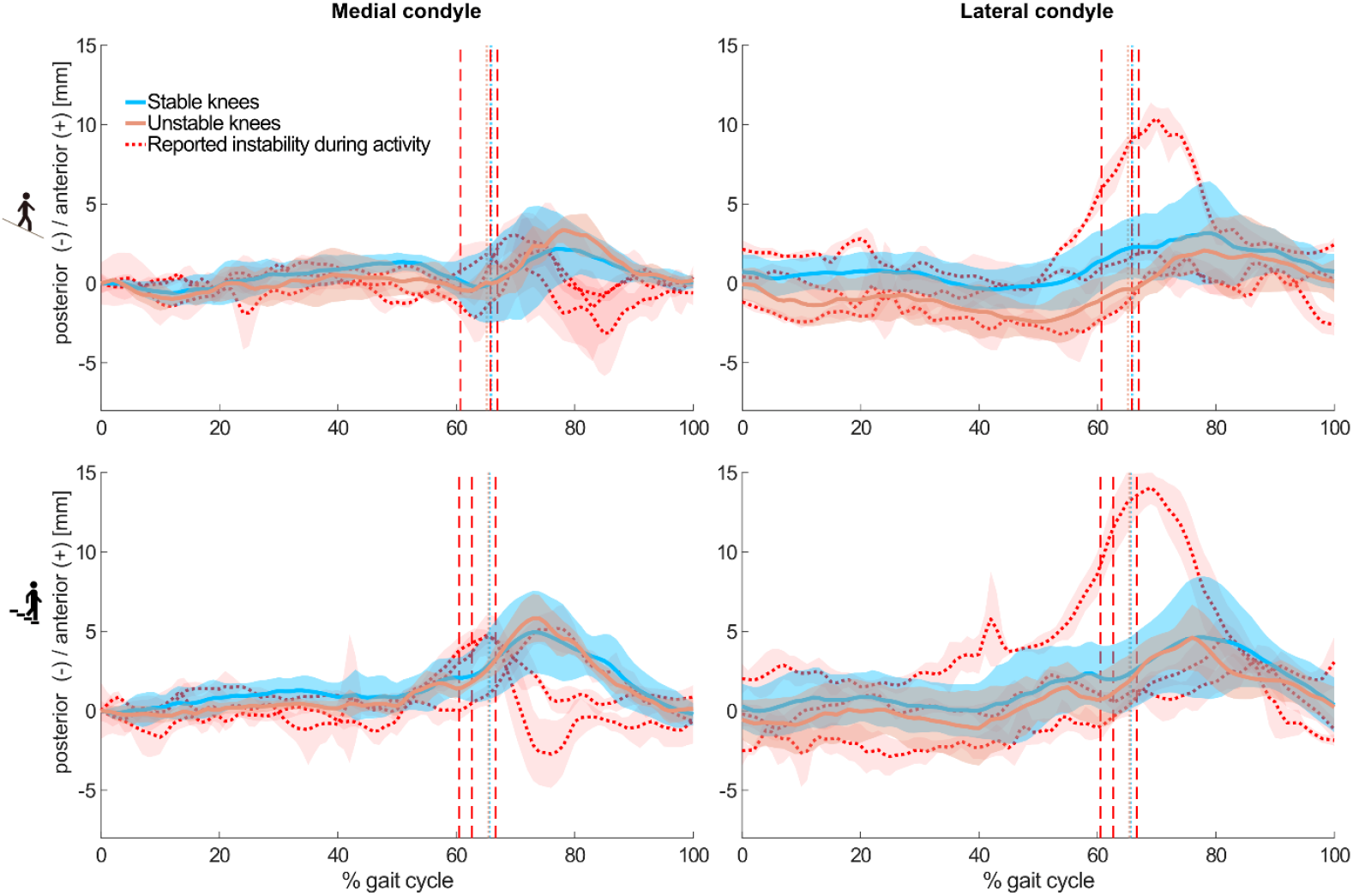
Mean and standard deviations (shaded) across subjects of the tibiofemoral A-P translations in the stable (8/8, blue), and unstable (5/8, orange) groups without reported instability during the measured activities. In addition, mean and standard deviations (shaded) across trials of three individuals from the unstable group who reported instability during the measured activities are shown in red. Dashed colour lines indicate the mean toe-offs for the stable and unstable groups, as well as for each unstable TKA knee with specifically reporting instability.

### Muscle synergy analysis

In general, a comparable number of muscle synergies were extracted between stable and unstable knees during level walking (stable 3.7±0.5 vs unstable 3.5±0.5), downhill walking (3.6±0.5 vs 3.6±0.5), and stair descent (3.4±0.5 vs 3.2±0.9). By rounding these number of synergies, the number of classified fundamental synergies was set to 4 for level walking and downhill walking, and 3 for stair descent (Figure 4). The ratio of classifiable muscle synergies to total muscle synergies was slightly increased from level gait to downhill walking in the stable group (89% vs 94%) but dropped from 95% to 80% in the unstable group. The ratios were lower (stable 78% vs unstable 77%) in both groups during stair descent. During level and downhill walking, each classified synergy was dominated by the knee extensor (quadriceps), plantarflexor (gastrocnemii), dorsiflexor (tibialis anterior), or knee flexor (hamstrings) muscle groups, individually. These 4 distinct synergies were observable among the stable knees in a comparable manner between level walking (90%, 100%, 60% and 90% of subjects exhibited this pattern) and downhill walking (100%, 90%, 60% and 90%). However, these synergies were less frequently observed in unstable knees during downhill walking (86%, 71%, 86%, and 43%) compared to level walking (100%, 100%, 50%, and 83%), as well as to the stable knees during downhill walking. During stair descent, 3 synergies, dominated by the knee extensors, dorsi- and knee-flexors, or plantarflexors accordingly, could be identified in both the stable and unstable cohorts, but were more prevalently found in the stable knees (100%, 90%, and 100%) than their unstable counterparts (100%, 75%, and 75%).

**Figure 4:**
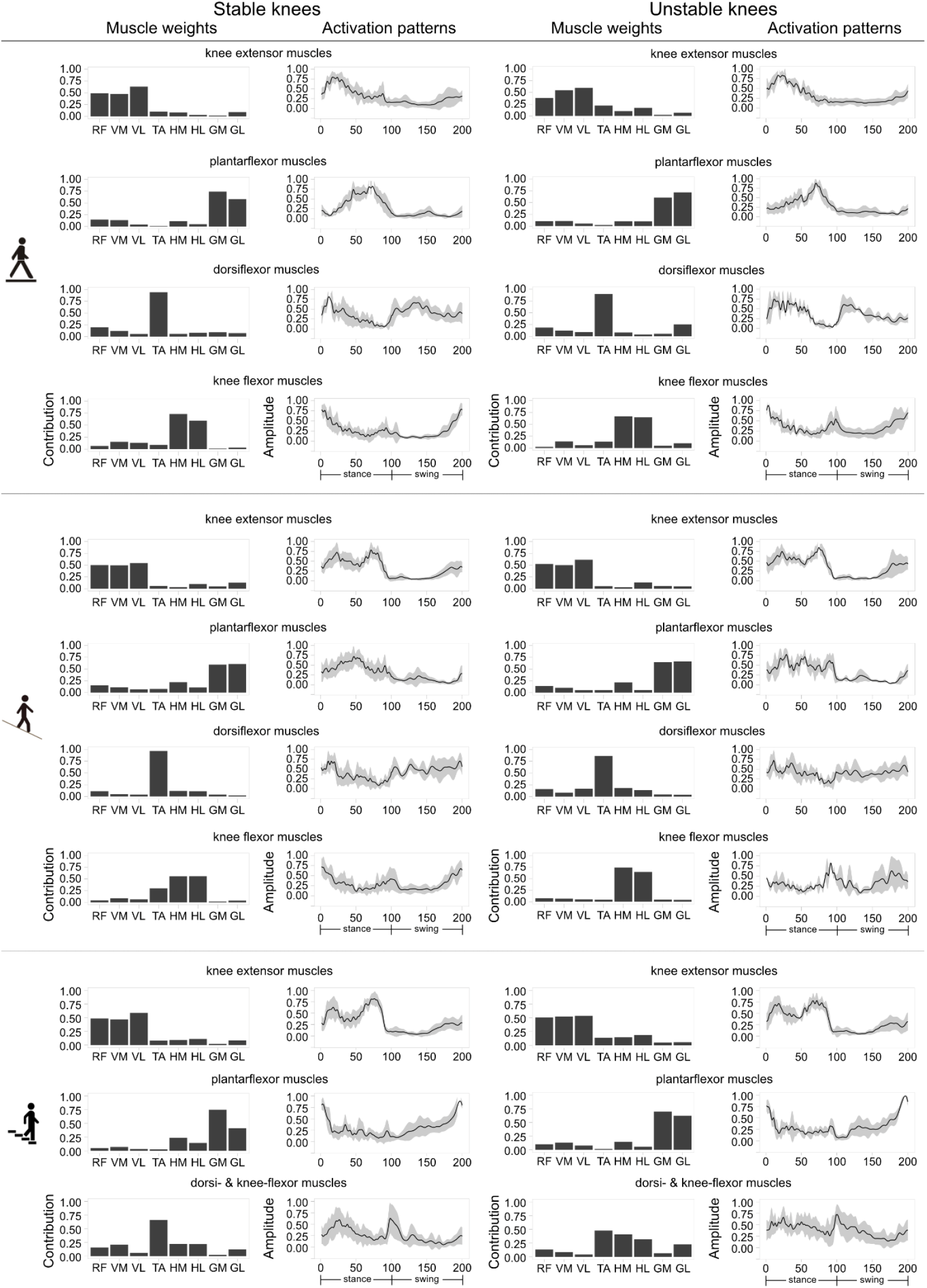
Classified muscle synergies in both stable and unstable TKA knees during level walking, downhill walking, and stair descent. Muscle synergy weights, as well as means (solid lines) and standard deviations (shaded areas) of the corresponding time/amplitude-normalized activation patterns are presented for each activity. RF: rectus femoris, VM: vastus medialis, VL: vastus lateralis, TA: tibialis anterior, HM: medial hamstrings, HL: lateral hamstrings, GM: gastrocnemius medialis, GL: gastrocnemius lateralis.

Muscle synergies exhibited phase-dependent activation patterns. Knee extensor muscles were predominantly activated during early stance (level walking) or throughout stance (downhill walking and stair descent) phases, while the plantarflexors contributed to the late stance (level walking), throughout stance (downhill walking), and mid-late swing (stair descent) phases (Figure 4). High contributions of the knee flexor muscles were identified during late swing/early stance (all activities), as well as at toe-off (downhill walking). As expected, the dorsiflexors exhibit a typical pattern of activation, showing strong contributions at early stance and early swing during level walking, while presenting a more sustained activation during downhill walking and stair descent.

The FWHM of the classified synergy activation patterns of the dorsi- and knee-flexors were significantly higher in the unstable than the stable knees (ES=1.49, p=0.01) during stair descent (Table 4). Comparable results were found in the FWHM of the knee extensor and plantarflexor activations between the two groups during all activities. No differences were observed in the CoA of any synergies except the activation pattern dominated by dorsi-flexors during level walking (ES=2.17, p<0.01) (Table 5), but the corresponding classified muscle synergy was only found in 3 out of the 8 unstable knees. The one-way ANOVA showed similar muscle weights between the groups during level walking and stair descent (Table S1)

**Table 4:** Mean±standard deviation of full width at half maximum (FWHM) of the synergistic activation patterns corresponding to knee extensor, plantarflexor, dorsiflexor, and knee flexor muscle groups during level walking, downhill walking, and stair descent. Significant differences were observed between stable and unstable knees during stair descent in dorsi- (^a^) and knee-flexor muscles (^b^), with effect size of 1.49. *The classified synergy corresponding to dorsi- and knee-flexor muscles was only observed in a small number of unstable knees (3/8).

| FWHM | Knee extensors |  |  | Plantarflexors |  |  | Dorsiflexors |  |  | Knee flexors |  |  |
| --- | --- | --- | --- | --- | --- | --- | --- | --- | --- | --- | --- | --- |
|  | stable | unstable | p | stable | unstable | p | stable | unstable | p | stable | unstable | p |
| Level walking | 29.3±8.1 | 28.6±13.1 | 0.55 | 24.3±7.9 | 25.5±6.7 | 0.38 | 37.4±13.2 | 27.7±8.3 | 0.11* | 21.7±6.2 | 30.4±9.5 | 0.06 |
| Downhill walking | 31.2±9.4 | 34.1±6.6 | 0.26 | 20.2±7.3 | 22.6±8.4 | 0.29 | 35.2±15.9 | 31.1±13.2 | 0.32 | 20.2±5.7 | 21.6±5.5 | 0.37* |
| Stair descent | 31.8±12.2 | 36.0±7.3 | 0.20 | 26.0±9.4 | 33.5±15.1 | 0.12 | 20.1±8.1 | 35.0±12.5 | 0.01<sup>a</sup> | 20.1±8.1 | 35.0±12.5 | 0.01<sup>b</sup> |

**Table 5:**
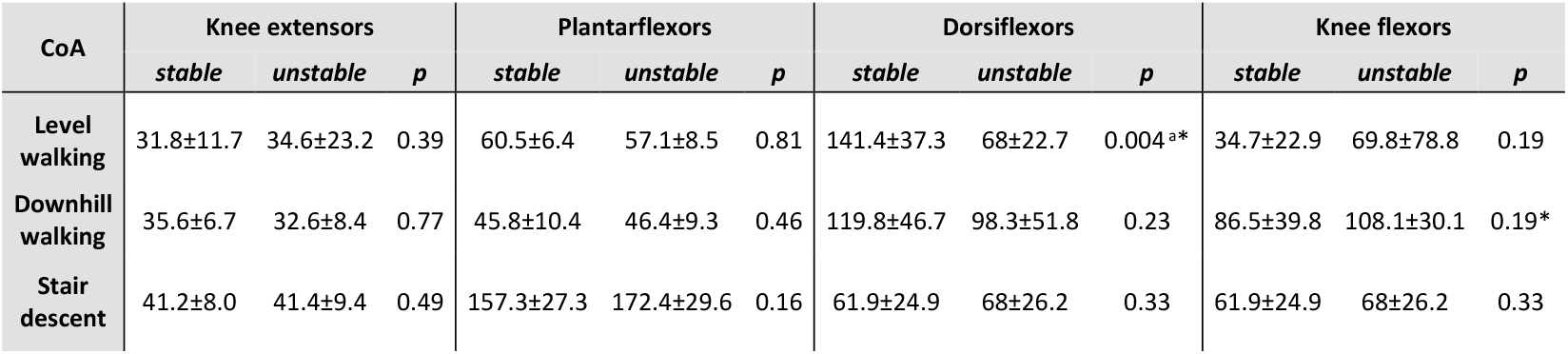
Mean±standard deviation of centre of activity (CoA) of the synergistic activation patterns corresponding to knee extensor, plantarflexor, dorsiflexor, and knee flexor muscle groups during level walking, downhill walking, and stair descent. A significant difference was observed between stable and unstable knees during stair descent in dorsiflexor muscles (^a^), with effect size of 2.17. *The classified synergy corresponding to dorsi- and knee-flexor muscles was only observed in a small number of unstable knees (3/8).

## Discussion

Knee instability is one of the most common reasons for unsatisfactory outcomes after TKA, accounting for up to 26% of revisions (4, 41). However, the relationships between self-reported assessment of instability and knee functionality in daily living remain unclear. Using moving video-fluoroscopy and EMG, we compared kinematic parameters of knee joint stability, as well as muscle synergy patterns, between subjects who had expressed concern at the stability of their knee, and those who were happy with its functional performance. Our results indicate that kinematic parameters including A-P translations and knee rotations, as well as their RoMs during functional activities, were generally comparable between stable and unstable TKA knees. However, increased heterogeneity in muscle synergy patterns between subjects and across activities, especially during challenging tasks such as stair descent, as well as prolonged activation of the classified knee flexor synergy, were observed in the unstable group. These differences between groups plausibly reveal long-lasting muscular adaptation strategies that develop as a compensation mechanism for feelings of joint instability and serve to prevent possible unstable events. Despite these modifications in muscular strategies, however, specific cases of acute instability were still reported by some subjects during the measurements, and analysis of their data revealed clear deviations in kinematic patterns from those of both the stable and unstable groups. These observations suggest that accurate movement analysis might be highly sensitive for detecting acute instability events, but might be less robust in identifying general joint instability.

The range of A-P translation in both groups studied here was comparable to previous studies during functional activities in good outcome TKAs with similar UC inlay geometries (28, 42, 43). The so-called ’paradoxical anterior translation ’ pattern that is commonly observed in tibiofemoral kinematics after TKA (44) was also observed in both groups during level walking and became even more evident during downhill walking and stair descent. Interestingly, while the stable group showed comparable A-P translation RoMs on both condyles, the unstable group exhibited medio-lateral differences during the stance phase of downhill walking and the swing phase of stair descent (Table 2). However, the small magnitudes of these differences are unlikely to be clinically relevant for identifying subjects with joint instability. Here, it should be noted that the assessment of subject-specific tibiofemoral kinematics was capable of identifying acute instability events. *In vivo* kinematics of three unstable knees who reported instability during downhill walking and stair descent showed specific A-P motion patterns, with an early, rapid condylar posterior translation followed by an additional anterior translation in the early/mid-swing phase (Figure 3). Such kinematic patterns stand out from those of both the stable cohort and even the unstable knees that did not report instability during the measured activities. Although our findings suggest that the accurate assessment of tibiofemoral kinematics has the potential to identify acute instability events, we have not been able to detect a common distinct kinematic pattern throughout the cohort of unstable knees.

Extensive efforts have been made to understand kinematic adaptations in knee pathologies including osteoarthritis (OA) and posterior cruciate ligament deficiency (PCLD). However, no significant differences have yet been found between PCLD and healthy knees in abduction/adduction or in internal/external rotation (45), nor in knee flexion/extension between OA knees and healthy controls (46) during level gait. During more challenging activities such as stair descent, the differences in knee flexion angle and extensor torque between PCLD and healthy knees have been found to be statistically significant but presented no outstanding clinical consequences (47). Interestingly, the structural deficits presented in such cases did not seem to result in clear kinematic deviations. Similarly in our study of knee joint instability, it seems that tibiofemoral movement patterns become well-compensated/controlled, plausibly through altered muscular control mechanisms.

It was previously found that unsatisfactory TKA knees present fewer muscle synergies for locomotion compared to satisfactory TKA knees and healthy controls (31). However, our results suggest that more heterogenous synergy patterns were developed in the unstable compared to the stable knees in the long term, albeit with a comparable number of synergies extracted from both groups in all activities. A higher number of non-classifiable, subject-specific muscle synergies was identified in the unstable compared to the stable group during level walking, and this number further increased during the more demanding activities of downhill walking and stair descent. This finding suggests that more challenging locomotor activities have a greater potential for provoking knee instability, which is plausibly linked to the requirement for a specific or individual response to each unstable situation or musculoskeletal deficit. Higher FWHM in the knee-/dorsi-flexor-controlled activation patterns were observed in the unstable knees compared to their stable counterparts during stair descent, which further indicates that the feeling of instability can permanently trigger prolonged activation of flexor muscle groups. This finding is in line with previous studies reporting the widening of muscle activation patterns in pathologies (39, 48, 49), early developmental stage (39, 50), ageing (51, 52) and presence of external mechanical perturbations (53). A possible explanation for this peculiar behaviour can be found in mouse studies, where similar temporal modifications of muscle activity have been associated with a lack of sensory feedback from proprioceptors (54-56). Interestingly, our results showed that knee extensor-flexor co-activation was present throughout the stance phase of the gait cycle in both TKA groups in all activities. However, this co-activation pattern was generally not reported in healthy stable knees (53, 57, 58). This finding shows that co-activation of knee extensors and flexors appears after TKA and persists in the long term. Moreover, we found a major involvement in TKA knees of knee flexors in the propulsion/early swing phase during all activities, contrary to what is typically found in healthy subjects (51, 53, 57, 58). These activation bursts of the knee flexors possibly serve as an additional adaptive strategy to ensure joint stability during daily activities (25, 59).

There are some limitations to this retrospective observational study. Firstly, only small sample sizes were considered. Since the study protocols (implant and inlay type, patient age etc) were all tightly controlled, however, we have been able to present well-standardised conditions for all subjects. Moreover, self-reported instability is highly subjective and activity-dependent, which is visible in the high inter-subject kinematic variability, and not all patients in the unstable group reported a feeling of instability during measurement. Our analyses have therefore not been able to capture all the characteristics of self-reported instability, but have rather been able to make progress in understanding the relationships between several important confounding factors, as well as the individual compensation mechanisms that have developed due to joint instability. The unstable knees in this study demonstrated highly heterogenous, unclassifiable muscle synergies with variable activation patterns, suggesting patient-specific muscular strategies. However, it remains unclear whether the observed synergy patterns are a hallmark for classifying knee stability. Lastly, only a single implant type was examined in this study. Here, we acknowledge that considerable further investigation would be required to establish whether the same or similar results are observed in subjects with other implants. Moreover, it is likely that more challenging tasks (e.g. stop-and-go activities) could expose the subjects to more frequent instability events than the daily activities investigated in our study, but such tasks are hard to standardise for reliable assessment of kinematic instability.

## Conclusion

This study quantitatively investigated the tibiofemoral kinematics and muscle synergy patterns in stable and unstable TKA knees during functional activities of daily living using moving video-fluoroscopy and EMG. Our results showed that condylar A-P translations, rotations, as well as their RoMs were comparable between stable and unstable TKA knees. However, subjects who reported instability events during measurement showed distinct, subject-specific tibiofemoral kinematic patterns that differed from those in stable knees. More importantly, the normal tibiofemoral kinematics observed in the unstable group were accompanied by a greater heterogeneity in muscle synergy patterns and prolonged activation of knee flexor muscles compared to the stable group. These differences between groups plausibly reveal long-lasting muscular adaptation strategies that develop as a compensation mechanism for feelings of joint instability and serve to prevent possible unstable events. Our findings suggest that the analysis of muscle synergies is able to identify muscular adaptation that likely results from feelings of joint instability – and is therefore indicative of underlying chronic knee instability – whereas tibiofemoral kinematics are sensitive for detecting acute instability events during functional activities.

## Data Availability

All data generated or analysed in this study are included in the manuscript and supplementary data files.

https://doi.org/10.3929/ethz-b-000584582

## Acknowledgements

The authors would like to thank Michael PlÜss, Samara Stulz, Dr. Barbara Postolka, Julia KÜndig, Alexander Mattmann and Peter Schwilch for supporting construction of the experimental set up, as well as the lab measurements. We are also grateful to Zimmer Biomet for providing the CAD models of the Persona implant components.

---

**Table S1:** Post-hoc pair-wise comparisons of one-way ANOVA results on hamstrings-dominant classified synergy module during stair descent. Bold values indicate the comparison of the same muscle between stable and unstable groups. RF: rectus femoris, VM: vastus medial, VL: vastus lateral, TA: tibialis anterior, HM: hamstrings medial, HL: hamstring lateral, GM: gastrocnemius medial, GL: gastrocnemius lateral.

| ANOVA | GL_stable | GM_stable | HL_stable | HM_stable | RF_stable | TA_stable | VL_stable | VM_stable |
| --- | --- | --- | --- | --- | --- | --- | --- | --- |
| GL_unstable | 0.658 | - | - | - | - | - | - | - |
| GM_unstable | - | 0.87007 | - | - | - | - | - | - |
| HL_unstable | - | - | 0.67617 | - | - | - | - | - |
| HM_unstable | - | - | - | 0.3568 | - | - | - | - |
| RF_unstable | - | - | - | - | 0.94069 | - | - | - |
| TA_unstable | - | - | - | - | - | 0.36318 | - | - |
| VL_unstable | - | - | - | - | - | - | 0.94945 | - |
| VM_unstable | - | - | - | - | - | - | - | 0.56587 |

## Notes

### Competing Interest Statement

The authors have declared no competing interest.

### Clinical Trial

BASEC no. 2019-01242

### Funding Statement

The authors would like to thank the Orthopaedics Hospital, Dongxiang, China, for financially supporting Longfeng Rao during his PhD.

### Author Declarations

Kantonale Ethikkommission Zurich Geschaftsfuhrer Stampfenbachstrasse 121 Postfach 8090 Zurich

## References

1. Hosseini Nasab SH, Smith CR, Schutz P, Postolka B, List R, Taylor WR. Elongation Patterns of the Collateral Ligaments After Total Knee Arthroplasty Are Dominated by the Knee Flexion Angle. Front Bioeng Biotechnol. 2019;2019:323.

2. Song SJ, Detch RC, Maloney WJ, Goodman SB, Huddleston JI. Causes of Instability After Total Knee Arthroplasty. Journal of Arthroplasty. 2014;29(2):360–4.

3. Nagle M, Glynn A. Midflexion Instability in Primary Total Knee Arthroplasty. J Knee Surg. 2020;33(5):459–65.

4. Parratte S, Pagnano MW. Instability after total knee arthroplasty. J Bone Joint Surg Am. 2008;90a(1):184–94.

5. Roos EM, Roos HP, Lohmander LS, Ekdahl C, Beynnon BD. Knee injury and osteoarthritis outcome score (KOOS) - Development of a self-administered outcome measure. J Orthop Sport Phys. 1998;28(2):88–96.

6. Dawson J, Fitzpatrick R, Murray D, Carr A. Questionnaire on the perceptions of patients about total knee replacement. J Bone Joint Surg Br. 1998;80b(1):63–9.

7. Zahiri CA, Schmalzried TP, Szuszczewicz ES, Amstutz HC. Assessing activity in joint replacement patients. J Arthroplasty. 1998;13(8):890–5.

8. Bellamy N, Buchanan WW, Goldsmith CH, Campbell J, Stitt LW. Validation study of WOMAC: a health status instrument for measuring clinically important patient relevant outcomes to antirheumatic drug therapy in patients with osteoarthritis of the hip or knee. J Rheumatol. 1988;15(12):1833–40.

9. Mears SC, Severin AC, Wang J, Thostenson J, Mannen EM, Stambough JB, et al. Inter-rater reliability of clinical testing for laxity after knee replacement. J Arthroplasty. 2022.

10. White SH, Oconnor JJ, Goodfellow JW. Sagittal Plane Laxity Following Knee Arthroplasty. J Bone Joint Surg Br. 1991;73(2):268–70.

11. Ishii Y, Matsuda Y, Ishii R, Sakata S, Omori G. Sagittal laxity in vivo after total knee arthroplasty. Arch Orthop Traum Su. 2005;125(4):249–53.

12. Murer M, Falkowski AL, Hirschmann A, Amsler F, Hirschmann MT. Threshold values for stress radiographs in unstable knees after total knee arthroplasty. Knee Surg Sports Traumatol Arthrosc. 2020.

13. Moser LB, Koch M, Hess S, Prabhakar P, Rasch H, Amsler F, et al. Stress Radiographs in the Posterior Drawer Position at 90° Flexion Should Be Used for the Evaluation of the PCL in CR TKA with Flexion Instability. J Clin Med. 2022;11(4):1013.

14. Jung TM, Reinhardt C, Scheffler SU, Weiler A. Stress radiography to measure posterior cruciate ligament insufficiency: a comparison of five different techniques. Knee Surg Sports Traumatol Arthrosc. 2006;14(11):1116–21.

15. Moewis P, Duda GN, Jung T, Heller MO, Boeth H, Kaptein B, et al. The Restoration of Passive Rotational Tibio-Femoral Laxity after Anterior Cruciate Ligament Reconstruction. PLoS One. 2016;11(7):e0159600.

16. Lorbach O, Brockmeyer M, Kieb M, Zerbe T, Pape D, Seil R. Objective measurement devices to assess static rotational knee laxity: focus on the Rotameter. Knee Surg Sports Traumatol Arthrosc. 2012;20(4):639–44.

17. Schuster AJ, von Roll AL, Pfluger D, Wyss T. Anteroposterior stability after posterior cruciate-retaining total knee arthroplasty. Knee Surg Sports Traumatol Arthrosc. 2011;19(7):1113–20.

18. Oh CS, Song EK, Seon JK, Ahn YS. The effect of flexion balance on functional outcomes in cruciate-retaining total knee arthroplasty. Arch Orthop Trauma Surg. 2015;135(3):401–6.

19. Jones DP, Locke C, Pennington J, Theis JC. The effect of sagittal laxity on function after posterior cruciate-retaining total knee replacement. J Arthroplasty. 2006;21(5):719–23.

20. Stacoff A, Diezi C, Luder G, Stussi E, Kramers-de Quervain IA. Ground reaction forces on stairs: effects of stair inclination and age. Gait Posture. 2005;21(1):24–38.

21. Simon JC, Della Valle CJ, Wimmer MA. Level and Downhill Walking to Assess Implant Functionality in Bicruciate- and Posterior Cruciate-Retaining Total Knee Arthroplasty. J Arthroplasty. 2018;33(9):2884–9.

22. Berman AT, Bosacco SJ, Israelite C. Evaluation of Total Knee Arthroplasty Using Isokinetic Testing. Clin Orthop Relat R. 1991(271):106–13.

23. Walsh M, Woodhouse LJ, Thomas SG, Finch E. Physical impairments and functional limitations: A comparison of individuals 1 year after total knee arthroplasty with control subjects. Phys Ther. 1998;78(3):248–58.

24. Lundberg HJ, Rojas IL, Foucher KC, Wimmer MA. Comparison of Antagonist Muscle Activity During Walking Between Total Knee Replacement and Control Subjects Using Unnormalized Electromyography. J Arthroplasty. 2016;31(6):1331–9.

25. Benedetti MG, Catani F, Bilotta TW, Marcacci M, Mariani E, Giannini S. Muscle activation pattern and gait biomechanics after total knee replacement. Clin Biomech. 2003;18(9):871–6.

26. Thomas AC, Judd DL, Davidson BS, Eckhoff DG, Stevens-Lapsley JE. Quadriceps/hamstrings coactivation increases early after total knee arthroplasty. Knee. 2014;21(6):1115–9.

27. Taylor WR, Schutz P, Bergmann G, List R, Postolka B, Hitz M, et al. A comprehensive assessment of the musculoskeletal system: The CAMS-Knee data set. J Biomech. 2017;2017:32–9.

28. Schutz P, Taylor WR, Postolka B, Fucentese SF, Koch PP, Freeman MAR, et al. Kinematic Evaluation of the GMK Sphere Implant During Gait Activities: A Dynamic Videofluoroscopy Study. J Orthop Res. 2019;37(11):2337–47.

29. Schutz P, Postolka B, Gerber H, Ferguson SJ, Taylor WR, List R. Knee implant kinematics are task-dependent. J R Soc Interface. 2019;16(151):20180678.

30. List R, Schutz P, Angst M, Ellenberger L, Datwyler K, Ferguson SJ, et al. Videofluoroscopic Evaluation of the Influence of a Gradually Reducing Femoral Radius on Joint Kinematics During Daily Activities in Total Knee Arthroplasty. J Arthroplasty. 2020;35(10):3010–30.

31. Ardestani MM, Malloy P, Nam D, Rosenberg AG, Wimmer MA. TKA patients with unsatisfying knee function show changes in neuromotor synergy pattern but not joint biomechanics. J Electromyogr Kinesiol. 2017;2017:90–100.

32. List R, Postolka B, Schutz P, Hitz M, Schwilch P, Gerber H, et al. A moving fluoroscope to capture tibiofemoral kinematics during complete cycles of free level and downhill walking as well as stair descent. PLoS One. 2017;12(10):e0185952.

33. Foresti M. In vivo measurement of total knee joint replacement kinematics and kinetics during stair descent: ETH; 2009.

34. Burckhardt K, Szekely G, Notzli H, Hodler J, Gerber C. Submillimeter measurement of cup migration in clinical standard radiographs. IEEE Trans Med Imaging. 2005;24(5):676–88.

35. Grood ES, Suntay WJ. A joint coordinate system for the clinical description of three-dimensional motions: application to the knee. J Biomech Eng. 1983;105(2):136–44.

36. Santuz A. musclesyneRgies: factorization of electromyographic data in R with sensible defaults. Journal of Open Source Software. 2022;7(74).

37. Santuz A, Ekizos A, Janshen L, Baltzopoulos V, Arampatzis A. On the Methodological Implications of Extracting Muscle Synergies from Human Locomotion. Int J Neural Syst. 2017;27(5):1750007.

38. Santuz A, Ekizos A, Janshen L, Mersmann F, Bohm S, Baltzopoulos V, et al. Modular Control of Human Movement During Running: An Open Access Data Set. Front Physiol. 2018;2018:1509.

39. Cappellini G, Ivanenko YP, Martino G, MacLellan MJ, Sacco A, Morelli D, et al. Immature Spinal Locomotor Output in Children with Cerebral Palsy. Front Physiol. 2016;2016:478.

40. Pataky TC, Robinson MA, Vanrenterghem J. Region-of-interest analyses of one-dimensional biomechanical trajectories: bridging 0D and 1D theory, augmenting statistical power. Peerj. 2016;4.

41. Wilson CJ, Theodoulou A, Damarell RA, Krishnan J. Knee instability as the primary cause of failure following Total Knee Arthroplasty (TKA): A systematic review on the patient, surgical and implant characteristics of revised TKA patients. Knee. 2017;24(6):1271–81.

42. Roberti di Sarsina T, Alesi D, Di Paolo S, Zinno R, Pizza N, Marcheggiani Muccioli GM, et al. In vivo kinematic comparison between an ultra-congruent and a posterior-stabilized total knee arthroplasty design by RSA. Knee Surg Sports Traumatol Arthrosc. 2021.

43. Cardinale U, Bragonzoni L, Bontempi M, Alesi D, Roberti di Sarsina T, Lo Presti M, et al. Knee kinematics after cruciate retaining highly congruent mobile bearing total knee arthroplasty: An in vivo dynamic RSA study. Knee. 2020;27(2):341–7.

44. Dennis DA, Komistek RD, Mahfouz MR. In vivo fluoroscopic analysis of fixed-bearing total knee replacements. Clin Orthop Relat Res. 2003(410):114–30.

45. Fontbote CA, Sell TC, Laudner KG, Haemmerle M, Allen CR, Margheritini F, et al. Neuromuscular and biomechanical adaptations of patients with isolated deficiency of the posterior cruciate ligament. Am J Sports Med. 2005;33(7):982–9.

46. Na A, Piva SR, Buchanan TS. Influences of knee osteoarthritis and walking difficulty on knee kinematics and kinetics. Gait Posture. 2018;2018:439–44.

47. Hooper D.M MMC, R. Crookenden, J. Ireland, J.P. Beacon. Gait adaptations in patients with chronic posterior instability of the knee. 2002.

48. Martino G, Ivanenko YP, d’Avella A, Serrao M, Ranavolo A, Draicchio F, et al. Neuromuscular adjustments of gait associated with unstable conditions. J Neurophysiol. 2015;114(5):2867–82.

49. Janshen L, Santuz A, Ekizos A, Arampatzis A. Fuzziness of muscle synergies in patients with multiple sclerosis indicates increased robustness of motor control during walking. Sci Rep. 2020;10(1):7249.

50. Dominici N, Ivanenko YP, Cappellini G, d’Avella A, Mondi V, Cicchese M, et al. Locomotor primitives in newborn babies and their development. Science. 2011;334(6058):997–9.

51. Santuz A, Brull L, Ekizos A, Schroll A, Eckardt N, Kibele A, et al. Neuromotor Dynamics of Human Locomotion in Challenging Settings. iScience. 2020;23(1):100796.

52. Dewolf AH, Sylos-Labini F, Cappellini G, Ivanenko Y, Lacquaniti F. Age-related changes in the neuromuscular control of forward and backward locomotion. PLoS One. 2021;16(2):e0246372.

53. Santuz A, Ekizos A, Eckardt N, Kibele A, Arampatzis A. Challenging human locomotion: stability and modular organisation in unsteady conditions. Sci Rep. 2018;8(1):2740.

54. Akay T, Tourtellotte WG, Arber S, Jessell TM. Degradation of mouse locomotor pattern in the absence of proprioceptive sensory feedback. Proc Natl Acad Sci U S A. 2014;111(47):16877–82.

55. Mayer WP, Akay T. Stumbling corrective reaction elicited by mechanical and electrical stimulation of the saphenous nerve in walking mice. J Exp Biol. 2018;221(Pt 13).

56. Santuz A, Akay T, Mayer WP, Wells TL, Schroll A, Arampatzis A. Modular organization of murine locomotor pattern in the presence and absence of sensory feedback from muscle spindles. J Physiol. 2019;597(12):3147–65.

57. Cappellini G, Ivanenko YP, Poppele RE, Lacquaniti F. Motor patterns in human walking and running. J Neurophysiol. 2006;95(6):3426–37.

58. Lacquaniti F, Ivanenko YP, Zago M. Patterned control of human locomotion. J Physiol. 2012;590(10):2189–99.

59. Hirokawa S. Muscular Co-Contraction and Control of Knee Stability. 1991.

